# Causes of false-negative rapid diagnostic tests for symptomatic malaria in the DRC

**DOI:** 10.1101/2020.08.25.20181354

**Authors:** Jonathan B. Parr, Eddy Kieto, Fernandine Phanzu, Paul Mansiangi, Kashamuka Mwandagalirwa, Nono Mvuama, Ange Landela, Joseph Atibu, Solange Umesumbu Efundu, Jean W. Olenga, Kyaw Lay Thwai, Camille E. Morgan, Madeline Denton, Alison Poffley, Jonathan J. Juliano, Pomie Mungala, Joris L. Likwela, Eric M. Sompwe, Antoinette K. Tshefu, Adrien N’Siala, Albert Kalonji

## Abstract

**Background:** The majority of *Plasmodium falciparum* malaria diagnoses in Africa are made using rapid diagnostic tests (RDTs) that detect histidine-rich protein 2. Increasing reports of false-negative RDT results due to parasites with deletions of the *pfhrp2* and/or *pfhrp3* genes (*pfhrp2/3*) raise concern about existing malaria diagnostic strategies. We previously identified *pfhrp2-*negative parasites among asymptomatic children in the Democratic Republic of the Congo (DRC), but their impact on diagnosis of symptomatic malaria is unknown.

**Methods:** We performed a cross-sectional study of false-negative RDTs in symptomatic subjects in 2017. Parasites were characterized by microscopy; RDT; *pfhrp2/3* genotyping and species-specific PCR assays; a multiplex bead-based immunoassay; and/or whole-genome sequencing.

**Results:** Among 3,627 symptomatic subjects, we identified 427 (11.8%) RDT-/microscopy+ cases. Parasites from eight (0.2%) samples were initially classified as putative *pfhrp2/3* deletions by PCR, but antigen testing and whole-genome sequencing confirmed the presence of intact genes. Malaria prevalence was high (57%) and non-falciparum co-infection common (15%). HRP2-based RDT performance was satisfactory and superior to microscopy.

**Conclusions:** Symptomatic malaria due to *pfhrp2/3*-deleted *P. falciparum* was not observed in the DRC.

Ongoing HRP2-based RDT use is appropriate for the detection of falciparum malaria in the DRC.

## BACKGROUND

Emergence of *Plasmodium falciparum* strains that evade detection by rapid diagnostic tests (RDTs) threatens progress toward malaria control and elimination in Africa.[1–3] These parasites have deletions involving the histidine-rich protein 2 and/or 3 (*pfhrp2/3*) genes, which encode the proteins detected by widely used RDTs throughout Africa. [4] Increasing reports of these parasites in select locations across Africa raise concern about the future of HRP2-based RDTs in the region. [5–7] Recent events in Eritrea, where a high frequency of false-negative RDTs due to these parasites triggered a change in national malaria diagnostic policy, emphasize the need for surveillance and a coordinated response to *pfhrp2/3-*deleted *P. falciparum.[8,9]*

We previously performed the first national survey of *pfhrp2* deletions and reported a 6.4% national prevalence of *pfhrp2-*negative parasites among asymptomatic children in the Democratic Republic of the Congo (DRC).[10] While this cross-sectional, nationally representative household survey enabled spatial analyses and initial population genetic analyses of *pfhrp2-*negative parasites in the DRC, it did not sample subjects with symptomatic malaria. In order to inform decisions about national malaria diagnostic testing policy, we undertook a cross-sectional survey of children and adults presenting to government health facilities in three provinces selected based on the prevalence of *pfhrp2-*negative parasites in our initial study. We hypothesized that *pfhrp2/3-*deleted parasites were responsible for missed clinical cases of falciparum malaria in the DRC.

Studies of *pfhrp2/3-*deleted *P. falciparum* are difficult due to the challenges of confirming the absence of these genes using conventional approaches. [11,12] These challenges are compounded by inconsistent laboratory methodologies across studies and inherent limitations of *pfhrp2/3* assays that can suffer from variable performance and cross-reactivity.[13,14] In addition, false-negative RDT results are common throughout Africa and typically caused by factors other than *pfhrp2/3* deletions, including operator error, lot-to-lot RDT variability, low-density infections beneath the RDT’s limit of detection, and infection by non-falciparum species. [15,16] In order to overcome these challenges, we performed a comprehensive molecular, serological, and genomic evaluation of symptomatic infections to define the causes of false-negative RDTs in the DRC and inform national diagnostic testing policy.

## RESULTS

### Study subjects

We enrolled 3,627 subjects with symptoms of malaria during November and December 2017, distributed across three provinces: 1,203 in Bas-Uele, 1,248 in Kinshasa, and 1,176 in Sud-Kivu (**Figure 1**). Study sites included 18 health facilities located in 18 distinct health areas, spanning six health zones (three health areas per health zone). Baseline characteristics of and malaria diagnostic testing results from enrolled subjects are displayed in **Table 1** and **Supplementary Table 1**. Age, gender, and the use of long-lasting insecticidal bednets was similar across health zones. Study sites included facilities with high, medium, and low symptomatic malaria prevalence. RDT-positive malaria, microscopy-positive malaria, and self-reported malaria diagnosis within the past six months were highest in Bas-Uele and lowest in Sud-Kivu.

**Figure 1.**
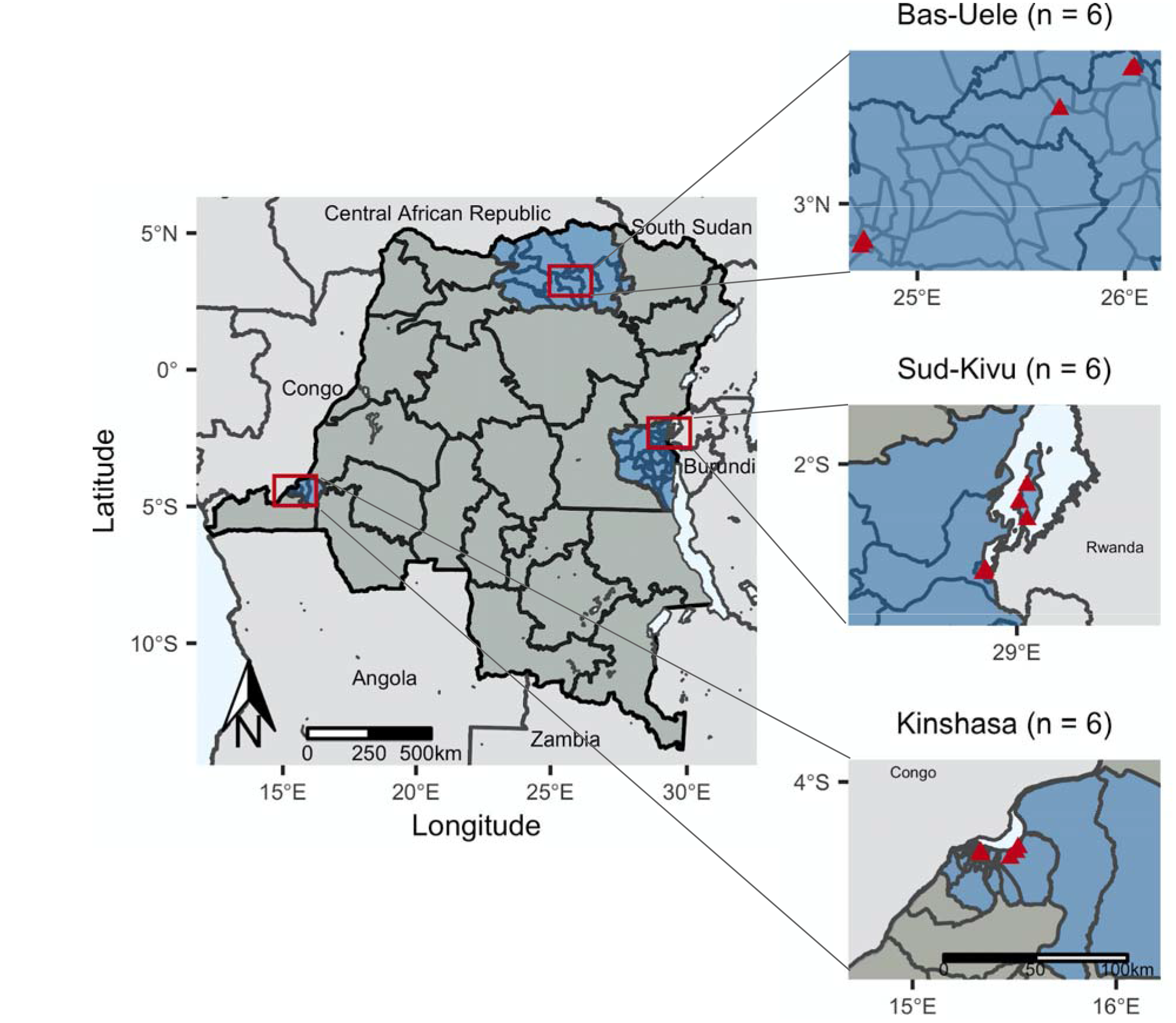
Study sites. included health facilities in eighteen health areas (triangles) in six health zones located within three provinces (n = number of facilities per province). Health areas in close proximity have overlapping points.

**Table 1.** Characteristics of enrolled study subjects.

|  | **Overall** | **Bas-Uele** | **Kinshasa** | **Sud-Kivu** |
| --- | --- | --- | --- | --- |
| Subjects, n | 3627 | 1203 | 1248 | 1176 |
| Health zones, n | 6 | 2 | 2 | 2 |
| Health areas, n | 18 | 6 | 6 | 6 |
| Age, mean years (SD) | 20.7 (18.5) | 19.5 (17.9) | 20.7 (18.7) | 22.0 (18.8) |
| Age strata, n (%) |  |  |  |  |
| <5 years | 1025 (28.7) | 370 (31.4) | 335 (26.8) | 320 (27.9) |
| 5-14 years | 579 (16.2) | 170 (14.4) | 271 (21.7) | 138 (12.0) |
| 15-24 years | 639 (17.9) | 232 (19.7) | 181 (14.5) | 226 (19.7) |
| 25-34 years | 513 (14.4) | 170 (14.4) | 144 (11.5) | 199 (17.4) |
| 35-44 years | 344 (9.6) | 101 (8.6) | 131 (10.5) | 112 (9.8) |
| 45-54 years | 249 (7.0) | 73 (6.2) | 108 (8.7) | 68 (5.9) |
| 55 years and older | 223 (6.2) | 62 (5.3) | 78 (6.2) | 83 (7.2) |
| Female gender, n (%) | 2130 (58.7) | 780 (64.8) | 646 (51.8) | 704 (59.9) |
| Pregnant, n (% of women) | 350 (16.4) | 105 (13.5) | 26 (4.0) | 219 (31.1) |
| Slept under bednet the night before, n (%) | 2238 (79.5) | 586 (75.9) | 902 (80.9) | 750 (80.9) |
| Diagnosed with malaria in the last six months, n (%) | 1556 (43.1) | 758 (63.0) | 462 (37.6) | 336 (28.6) |
| Microscopy-positive, n (%) | 1397 (38.7) | 500 (41.7) | 533 (43.0) | 364 (31.0) |
| RDT-positive, n (%) | 1545 (42.6) | 758 (63.0) | 380 (30.4) | 407 (34.6) |
| RDT-negative, microscopy-positive, n (%) | 426 (11.8) | 51 (4.2) | 267 (21.5) | 108 (9.2) |
| Parasites/μL by microscopy, geometric mean (geometric mean SD factor) | 2877 (6.9) | 3739 (6.5) | 1789 (7.2) | 4001 (6.3) |

### False-negative RDTs by microscopy

Among 3,627 subjects tested, 1,545 (42.6%) were RDT-positive and 1,397 (38.7%) were microscopy-positive, with 426 (11.8%) RDT-negative but microscopy-positive (‘false-negative RDT’). False-negative RDT results were more likely to occur at low microscopy parasite densities, with geometric means of 959 (geometric standard deviation [SD] factor 4.2) and 4,675 (geom. SD factor 6.9) parasites/μL for RDT-versus RDT+ samples, respectively (t-test p <0.001, **Figure 2**). We observed higher false-negative RDT prevalence in Kinshasa and Sud-Kivu than Bas-Uele, a pattern similar to RDT-/PCR+ results we observed in our original *pfhrp2* survey in asymptomatic children as part of the 2013-14 DRC DHS.[10] When tested by *P. falciparum lactate dehydrogenase* (*pfldh*) PCR, 368 (86%) of the 426 RDT-negative/microscopy-positive samples were PCR-negative, a finding consistent with false-positive microscopy calls in this cohort.

**Figure 2.**
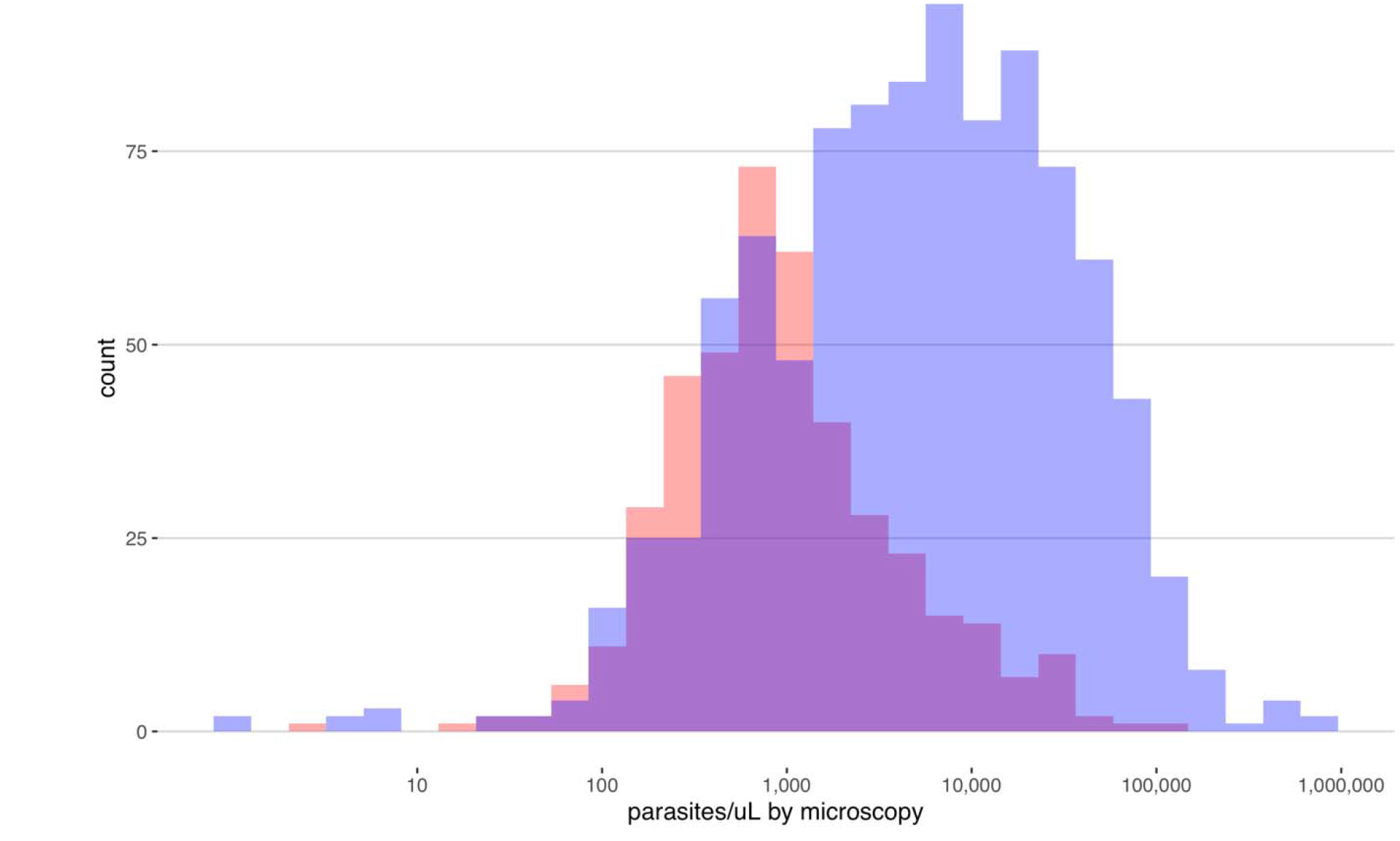
Parasite densities by microscopy among HRP2-based RDT-negative (red) and RDT-positive (blue) isolates. False-negative RDT results were more likely to occur at low microscopy parasite densities, with geometric means of 959 (geometric standard deviation [SD] factor 4.2) and 4,675 (geom. SD factor 6.9) parasites/μL for RDT-negative versus RDT-positive samples, respectively (t-test p <0.001).

### Pfhrp2/3 deletion genotyping by PCR

We performed *pfhrp2/3* genotyping using PCR on a subset of samples, including those collected from all 426 subjects with RDT-/microscopy+ results and from 429 RDT+/microscopy+ controls selected at random from the same province (**Supplementary Figure 1**). Among the RDT-samples, only 23 had parasite densities sufficient for *pfhrp2/3* deletion genotyping by *pfldh* qPCR (≥40 parasites/μL).[13] We further characterized these samples and 74 RDT-positive controls selected from the same facilities (n=97 total) using a series of PCR assays for *pfhrp2* and *pfhrp3*, and a final confirmatory PCR assay for P. *falciparum beta-tubulin* (*PfBtubulin*). Eight parasites were PCR-negative for *pfhrp2* or *pfhrp3* in duplicate despite having parasite densities well above the PCR assays’ limits of detection and successful amplification of a second single-copy gene, consistent with *pfhrp2/3* gene deletions using conventional PCR genotyping criteria. [4,5,17] PCR genotyping suggested six deletions among symptomatic RDT-/microscopy+ subjects (five *pfhrp2-/3-* and one *pfhrp2-/3+*) and two among RDT+/microscopy+ subjects (one *pfhrp2-/3+* and one *pfhrp2+/3-*). Parasite densities for these eight putative *pfhrp2/3-*deleted samples ranged from 84 to 102,700 parasites/μL by qPCR (median 2,929, interquartile range 1,314 to 4,572).

### Whole-genome sequencing of candidate pfhrp2/3 deletions

However, whole-genome sequencing (WGS) confirmed that all eight putative *pfhrp2/3-*deleted samples had parasites with intact *pfhrp2* and *pfhrp3* genes (**Figure 3**). All eight samples had >5-fold coverage across >80% of the genome, with median sequencing depth ranging from 66-254 reads/position (**Supplementary Figure 2**). Regions of reduced sequencing depth corresponded to differences in the number of histidine repeats compared to the 3D7 reference sequence and did not introduce frame-shift mutations. Mutations in PCR primer binding sites were not observed.

**Figure 3.**
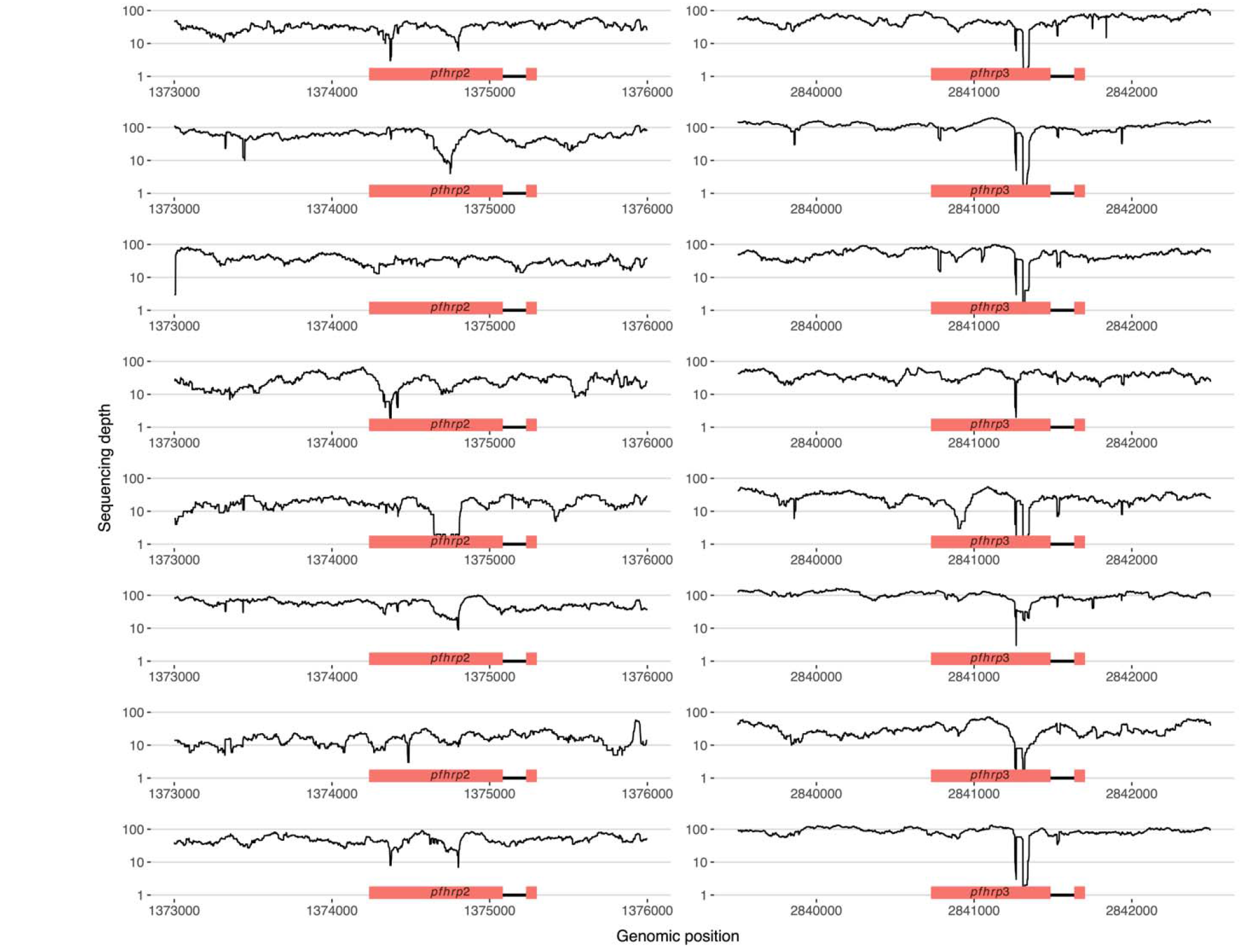
No *pfhrp2* or *pfhrp3* deletions were identified by whole-genome sequencing. WGS of five *pfhrp2-/3-*, two *pfhrp2-/3+*, and one *pfhrp2+/3-*parasites originally identified by PCR confirmed intact genes. Each row represents a distinct sample.

### Antigenemia assessment using a bead-based immunoassay

Luminex-based serological assessment further confirmed that all eight *pfhrp2/3-*PCR-negative samples had detectable HRP2 antigenemia, providing confidence that the intact genes observed during whole-genome sequencing encoded functional HRP2 and/or *P. falciparum* histidine-rich protein 3 (HRP3) proteins. We assessed HRP2 antigenemia in all 97 samples that had undergone *pfhrp2/3* genotyping by PCR. Comparing RDT-negative versus RDT-positive samples, positivity and background-subtracted mean fluorescence intensity (MFI) was similar between groups for all three antigens tested: HRP2, *Plasmodium* parasite lactate dehydrogenase (pLDH), and *Plasmodium* parasite aldolase (pAldolase) (**Table 2**). Surprisingly, the majority of RDT-samples tested had high levels of circulating HRP2 detected by Luminex, suggesting that the negative RDT results were likely due to operator error or RDT failure (**Figure 4**).

**Table 2.** Luminex bead-based immunoassay results. Frequencies and log-transformed mean values were compared using the Fisher’s exact test and t-test, respectively. Abbreviations: RDT, rapid diagnostic test; MFI-background, mean fluorescence intensity minus background; SD, standard deviation.

|  | Overall | RDT-negative | RDT-positive | p |
| --- | --- | --- | --- | --- |
| Tested, n | 97 | 23 | 74 |  |
| HRP2-positive, n (%) | 96 (99.0) | 22 (95.7) | 74 (100.0) | 0.24 |
| HRP2, MFI-background geom. mean (SD factor) | 10276 (2.9) | 11751 (1.7) | 6675 (7) | 0.31 |
| pLDH-positive, n (%) | 72 (74.2) | 15 (65.2) | 57 (77.0) | 0.28 |
| pLDH, MFI-background geom. mean (SD factor) | 408 (8.4) | 440 (9) | 319 (6.5) | 0.50 |
| pAldolase-positive, n (%) | 92 (94.8) | 21 (91.3) | 71 (95.9) | 0.56 |
| pAldolase, MFI-background geom. mean (SD factor) | 1419 (5.5) | 1571 (5.4) | 1022 (5.9) | 0.48 |

**Figure 4.**
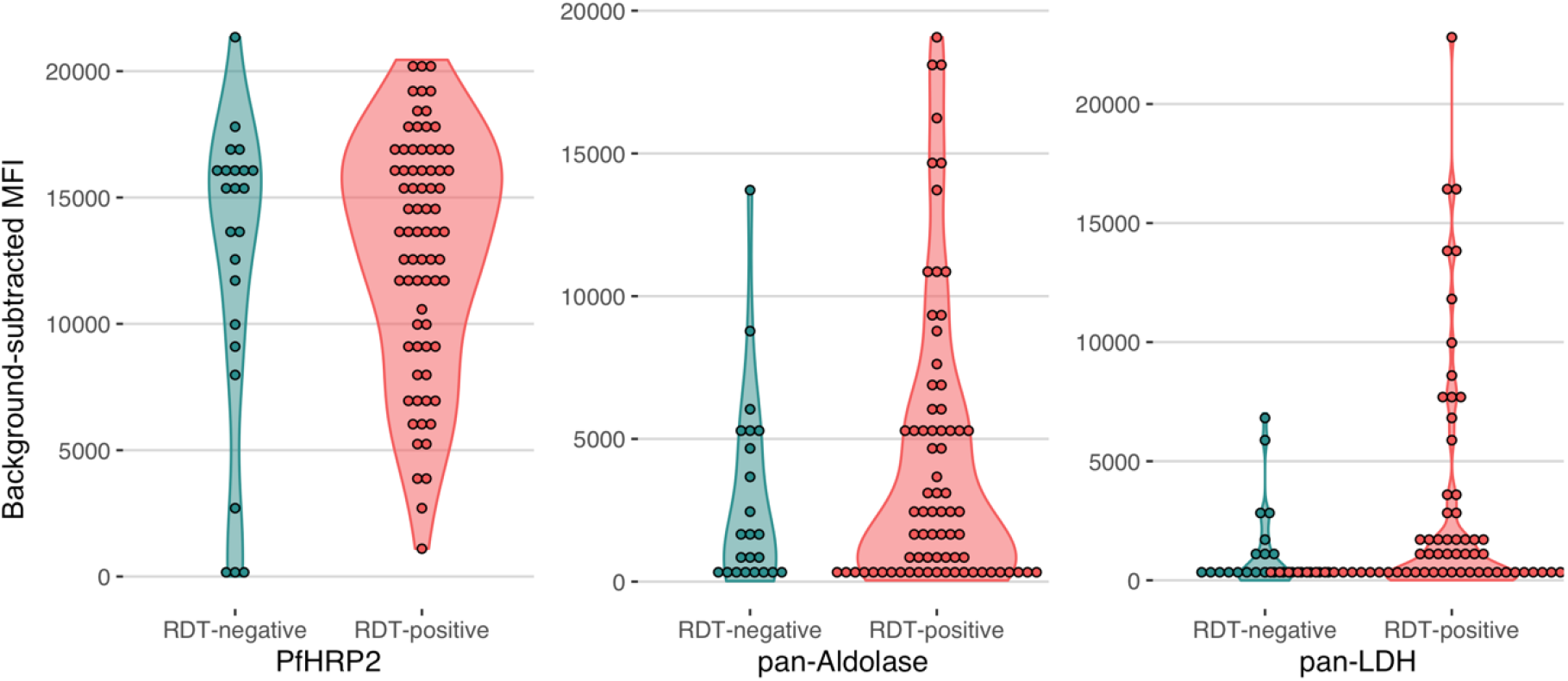
False-negative RDT results occurred in the setting of high HRP2 antigenemia. A Luminex bead-based immunoassay for three parasite antigens confirmed circulating HRP2 antigen in the majority of RDT-negative but PCR-positive/microscopy-positive isolates tested (RDT-negative). RDT-positive controls selected from the same facility are included for comparison. Abbreviations: MFI, mean fluorescence intensity; RDT, rapid diagnostic test.

### Non-falciparum malaria

Non-falciparum malaria is expected to cause HRP2-RDT-negative/microscopy-positive results and was common in our study cohort. Among 1,000 randomly selected samples that underwent species-identification using a series of real-time PCR assays (**Supplementary Figure 1**), malaria prevalence was high (57%), and non-falciparum co-infection with *P. falciparum* was common (15%, n=150) (**Table 3**). However, only 2.1% (n=11) of symptomatic cases were due to non-falciparum infections alone. *P. ovale* was observed in 12.0% (n=68) of symptomatic cases. Among the four (0.8%) symptomatic cases involving *P. vivax*, half involved *P. falciparum* and all were low density (<5 parasites/μL by semiquantitative 18S rRNA PCR). The majority of symptomatic P. *malariae* infections (86.9%, n=20/23) occurred as part of mixed infections with *P. falciparum* (**Supplementary Figure 3**). We were unable to determine the species in 19 samples that were positive by the pan-species 18S PCR assay in duplicate; all had negative P. *knowlesi* PCR results.

**Table 3.** Species identification by PCR among subjects with symptomatic malaria. Results of *Plasmodium* genus and species-specific 18S rRNA real-time PCR assays. Kruskal-Wallis p values are included for province-wise comparisons. Abbreviations: Pf, P. *falciparum;* Pm, P. *malariae;* Po, P. *ovale;* Pv, P. *vivax*.

|  | Overall | Bas-Uele | Kinshasa | Sud-Kivu | p |
| --- | --- | --- | --- | --- | --- |
| n | 1000 | 328 | 353 | 319 |  |
| *Plasmodium* (any species) PCR-positive, n (%) | 568 (56.8) | 280 (85.4) | 114 (32.3) | 174 (54.5) | <0.001 |
| *P. falciparum* PCR-positive, n (%) | 538 (53.8) | 268 (81.7) | 104 (29.5) | 166 (52.0) | <0.001 |
| Species identification by PCR, n (%) |  |  |  |  | 0.001 |
| *P. falciparum* only | 463 (81.5) | 210 (75.0) | 97 (85.1) | 156 (89.7) |  |
| *P. malariae* only | 2 (0.4) | 2 (0.7) | 0 (0.0) | 0 (0.0) |  |
| *P. ovale* only | 6 (1.1) | 1 (0.4) | 4 (3.5) | 1 (0.6) |  |
| *P. vivax* only | 2 (0.4) | 1 (0.4) | 0 (0.0) | 1 (0.6) |  |
| Mixed Pf and Pm | 16 (2.8) | 12 (4.3) | 1 (0.9) | 3 (1.7) |  |
| Mixed Pf and Po | 53 (9.3) | 42 (15.0) | 5 (4.4) | 6 (3.4) |  |
| Mixed Pf and Pv | 2 (0.4) | 1 (0.4) | 0 (0.0) | 1 (0.6) |  |
| Mixed Pm and Po | 1 (0.2) | 1 (0.4) | 0 (0.0) | 0 (0.0) |  |
| Mixed Pf, Pm, Po | 4 (0.7) | 3 (1.1) | 1 (0.9) | 0 (0.0) |  |
| *Plasmodium* positive, species undetermined | 19 (3.3) | 7 (2.5) | 6 (5.3) | 6 (3.4) |  |

### RDT performance

Assessment of RDT performance versus PCR suggested that false-negative RDT results in our cohort were commonly caused by RDT failure or operator error rather than parasite factors. Among the random subset of 1,000 samples that underwent 18S rRNA testing for all species, 134 (13.4%) of 538 *P. falciparum* 18S real-time PCR-positive samples were RDT-negative. RDT performance varied by province, with a larger proportion of RDT-/PCR+ results in provinces with higher *P. falciparum* prevalence by 18S rRNA PCR: Bas-Uele (19%), followed by Sud-Kivu (17%), and finally Kinshasa (5%) (**Supplementary Table 2**). Only a small proportion of samples (3.6%, n=36) were RDT+/PCR-, a finding not unexpected and suggestive of persistent PfHRP2 antigenemia after recent clearance of parasitemia.[18] When compared with PCR, RDTs were 75% sensitive and 92% specific, with good agreement (kappa = 0.66). Microscopy was 53% sensitive and 81% specific, with fair agreement with PCR (kappa = 0.33). Parasite densities as determined by microscopy and *pfldh* quantitative PCR (qPCR) had moderate correlation (Spearman correlation coefficient = 0.63, p <0.001, **Supplementary Figure 4**).

## DISCUSSION

We did not observe symptomatic malaria due to *pfhrp2-* or *pfhrp3*-deleted *P. falciparum* in this large, cross-sectional survey across three geographically disparate DRC provinces. The majority of RDT-negative/microscopy-positive results occurred in the setting of low or absent parasitemia. This finding implicates low parasite densities beneath the RDT’s limit of detection and false-positive microscopy results as the primary causes of RDT-microscopy discordance in the present study. Further assessment of RDT performance using microscopy, genus- and species-specific real-time PCR assays, and Luminex-based antigenemia assessment confirmed that RDT failure and/or user error also caused false-negative RDTs in the present study. However, the overall performance of HRP2-based RDTs was superior to microscopy and in good agreement with PCR.

These findings support continued use of HRP2-based RDTs in the DRC. They also contrast with the results of our prior study of asymptomatic children enrolled in the 2013-14 Demographic and Health Survey (DHS). There are several possible explanations for these differences. The present study enrolled symptomatic subjects in order to directly inform policy decisions about malaria case management. This study design could have inhibited our ability to identify *pfhrp2/3-*deleted parasites. We and others have proposed the hypothesis that parasites with deletions of the *pfhrp2* and/or *pfhrp3* genes and their flanking regions may be less fit,[6,10,19] and therefore less likely to cause symptomatic disease. Direct assessment of this hypothesis has not yet been performed *in vivo* or *in vitro*, to our knowledge. However, genetic cross experiments of the 3D7 (wild-type), DD2 (p/ftrp2-deleted), and HB3 (*pfhrp3-de\eted*) lab strains did not provide clear evidence of a fitness cost associated with deletion of either gene.[20,21] In addition, reports from Eritrea confirm that *pfhrp2/3-*deleted parasites can cause symptomatic and sometimes severe disease. [8]

Exhaustive analysis of putative *pfhrp2/3-*deleted parasites was needed to discern the status of both genes. The use of rigorous parasite density thresholds well above the downstream *pfhrp2/3* PCR assays’ limit of detection, [13] confirmation of successful amplification of multiple single-copy genes, and adherence to commonly accepted criteria[5] reduced the risk of inappropriate *pfhrp2/3* deletion calls. Only eight of 426 (1.8%) RDT-negative/microscopy-positive samples were identified as putative *pfhrp2/3* deletions during initial testing. However, we subsequently confirmed HRP2 antigenemia and intact *pfhrp2* and *pfhrp3* genes in all eight samples using highly sensitive antigen detection methods and WGS, respectively, confirming that these putative *pfhrp2/3-*deleted parasites were misclassified during initial testing.

These findings emphasize the challenges of confirming *pfhrp2/3* gene deletions and support the argument that a portion of *pfhrp2/3* deletion calls in our original study of asymptomatic children in the DRC were artifactual.[10,11] Even complex laboratory workflows conducted in accordance with commonly used deletion classification criteria are not always sufficient to eliminate the risk of misclassification of *pfhrp2/3* deletions. The use of advanced serological and next-generation sequencing methods improved the quality of our *pfhrp2/3* deletion assessment, allowed for a more robust evaluation of RDT performance, and enabled visualization of the genetic structure of the *pfhrp2* and *pfhrp3* genes and their flanking regions. While these methodologies are not widely available in resource-limited settings, they are now accessible through a network of laboratories that collaborate with the World Health Organization to support *pfhrp2/3* deletion surveillance[22] and in select locales in sub-Saharan Africa with advanced laboratory capacity.

Symptomatic malaria due non-falciparum species was common but usually occurred as part of mixed infections with *P. falciparum*. Although non-falciparum species are not detected by widely deployed HRP2-based RDTs, co-infection with *P. falciparum* is expected to trigger a positive RDT result and prompt treatment with artemisinin-combination therapy according to current DRC guidelines. Therefore, complications due to untreated symptomatic, non-falciparum malaria are likely uncommon, although the risk of relapse by P. *vivax* or P. *ovale* without proper diagnosis and terminal prophylaxis remains. Our findings are generally in-line with prior reports of non-falciparum infection among asymptomatic subjects in the DRC.[23–25]

Strengths of this study include its geographically diverse sampling locations, robust pipeline of conventional and advanced laboratory methodologies, and relevance to malaria case management. Indeed, these findings directly informed the DRC national malaria control program’s decision to continue the use of HRP2-based RDTs, despite evidence of *pfhrp2*-negative parasites from our initial study of asymptomatic subjects. Our experience in the DRC confirms the importance of basing policy decisions on careful studies of the target population - individuals presenting to health facilities with symptomatic malaria - rather than convenience sampling.

Limitations include our inability to discriminate *pfhrp2/3*-deleted from *pfhrp2/3-*intact strains in individuals infected by multiple *P. falciparum* strains. Neither the conventional methods nor the advanced Luminex-based HRP2 antigenemia assessment and WGS methods employed here are well-suited to identify gene deletions in mixed infections. Recently developed multiplexed qPCR methods[26] and amplicon-based deep sequencing approaches[2 7] have potential to elucidate *pfhrp2/3*-deleted minor variants in future large-scale surveys. Additionally, we restricted our *pfhrp2/3* deletion analysis to samples with ≥40 parasites/μL. This requirement was necessary to reduce the risk of misclassification due to DNA concentrations beneath the *pfhrp2/3* PCR assays’ limits of detection, [13] but it prevents us from commenting on the prevalence of deletions in lower density infections. Finally, this study was restricted to three provinces. These provinces spanned a range of malaria prevalence, but they do not capture the full diversity of the DRC, which is Africa’s second largest country by land mass and neighbors nine other countries.

In conclusion, ongoing HRP2-based RDT use is appropriate in the DRC. False-negative RDT results due to *pfhrp2/3* deletions were not observed among symptomatic subjects. Non-falciparum infection was an uncommon cause of false-negative results in the DRC, and RDT performance was superior to microscopy. Careful laboratory workflows are required during *pfhrp2/3* gene deletion analyses. Advanced serological and next-generation sequencing approaches can be used to improve the rigor and reproducibility of *pfhrp2/3* deletion surveillance efforts and to inform malaria diagnostic testing policy.

## METHODS

### Study population

We conducted a cross-sectional study of subjects presenting to hospitals and health centers across three provinces (Kinshasa, Bas-Uele, and Sud-Kivu) with symptoms of malaria. These provinces were selected based on results from our prior survey of asymptomatic children in the 2013-14 DHS and included both high-(Kinshasa, Sud-Kivu) and low-prevalence (Bas-Uele) of *pfhrp2*-negative parasites.[10] Two health zones were selected from each province, including one urban and one rural zone per province. Within each health zone, one general reference hospital and two health centers were selected as study sites, yielding six study sites per province and 18 study sites in total. Subjects of all ages were eligible for enrollment.

### Study procedures

Informed consent/assent was obtained from all study subjects prior to enrollment. All subjects received malaria RDT testing and treatment according to DRC national guidelines. Subjects underwent a study questionnaire and finger- or heel-prick whole blood collection for diagnostic testing by RDT and microscopy and DBS collection. RDT testing was performed using the WHO-prequalified, HRP2-based SD BIOLINE Malaria Ag P.f. (05FK50, Alere, Waltham, MA) according to manufacturer instructions. Thick-smear microscopy slides were read in the field, and thin smears fixed and transported to the National AIDS Control (PNLS) reference laboratory for confirmation and determination of parasite density. All thin smears were read by two microscopists, with discrepancies resolved by a third reader. DBS (Whatmann 903 Protein Saver cards, GE Healthcare Life Sciences, Marlborough, MA) were allowed to air dry at ambient temperature in the field, and stored in individual ziplock bags with desiccant at −20°C prior to and after shipment to the University of North Carolina at Chapel Hill for further testing. This study was approved by the Ethical Committee of the Kinshasa School of Public Health and the UNC Institutional Review Board.

### Pfhrp2/3 genotyping by PCR

DNA was extracted from DBS samples using Chelex and saponin. [28] All microscopy-positive, RDT-negative samples, in addition to an equal number of microscopy-positive, RDT-positive controls from each province were subjected to qPCR testing targeting the single-copy P. *pfldh* gene.[29] *Pfhrp2* and *pfhrp3* PCR genotyping was performed as previously described,[13] using conventional single-step *pfhrp2/3* PCR assays and a qualitative real-time PCR assay targeting the single-copy *PfBtubulin* gene (Supplementary File).[30-33] Only samples with ≥40 parasites/μL by qPCR (≥10-fold higher concentration than the *pfhrp2* and *pfhrp3* assays’ limits of detection) were subjected to *pfhrp2* and *pfhrp3* PCR to reduce the risk of misclassification of deletions. [13] Microscopy-positive, RDT-positive controls with ≥40 parasites/μL by qPCR were randomly selected from the same facility for *pfhrp2/3* genotyping. Samples were called *pfhrp2/3-*negative if they had ≥40 parasites/μL by *pfldh* qPCR, their *pfhrp2* and/or *pfhrp3* PCR assays were negative in duplicate, and they had successful amplification of *PfBtubulin* during a final confirmatory assay.

### Whole-genome sequencing

All *pfhrp2/3-*negative samples identified during initial testing were further assessed using whole-genome sequencing. DNA from these samples was enriched for *P. falciparum* prior to library prep using selective whole-genome amplification (sWGA) as previously described. [34] In brief, two sWGA reactions were performed in parallel, one using a custom primer set designed in our lab (JP9) and another using a primer set designed by Oyola *et al* (Probe_10).[35] sWGA products were pooled in equal volumes and acoustically sheared using a Covaris E220 instrument prior to library preparation using the Kapa HyperPrep kit (Roche Sequencing, Pleasanton, CA). Indexed libraries were pooled and sequenced at the UNC High Throughput Sequencing Facility using the HiSeq 2500 platform (Illumina, San Diego, CA) with 150bp, paired-end chemistry. Sequence reads were deposited into the Sequence Read Archive (accession numbers: pending).

### Evaluation for pfhrp2/3 deletions using whole-genome sequencing

Adapter sequences were trimmed from raw, paired sequence reads using *trimmomatic* and aligned to the *P. falciparum* 3D7 reference genome (PlasmoDB version 13.0) using *bwa mem* with default parameters.[36,37] Duplicates were marked and mate-pair information corrected using Picard Tool’s *MarkDupIicates* and *FixMatelnformation* functions, respectively. [3 8] Candidate indels were identified and realigned using GATK’s *RealignerTargetCreator* and *IndelAligner* functions, respectively. [39] Genome coverage was calculated using bedtool’s *genomecov* function and visualized using *ggplot2* in R (R Core Team, Vienna, Austria). [40,41] *Pfhrp2/3* deletions were called by visualization of aligned reads using the Integrative Genomics Viewer (Broad Institute, Cambridge, MA) and assessment of sequencing depth across the *pfhrp2/3* genes and their flanking regions. [42]

### Antigenemia assessment by Luminex

All DBS samples subjected to *pfhrp2/3* genotyping by PCR were also assayed for the following *Plasmodium* antigens: *Plasmodium* genus-specific aldolase (pAldolase) and lactate dehydrogenase (pLDH), as well as *P. falciparum* HRP2 by a bead-based multiplex assay as previously described.[43] Samples were assayed at 1:20 whole-blood concentration after elution from filter paper. Thresholds for antigen positivity for the three targets were determined by assaying 92 blood samples from US resident blood donors without history of international travel and determining mean and standard deviation of assay signal from this sample set. The mean plus three standard deviations of this sample set was used as the antigen positivity threshold.

### Non-falciparum assays

We used R to randomly select 1,000 samples for PCR-based species identification. DNA from these samples was first subjected to a pan*-Plasmodium* real-time PCR assay targeting the 18S rRNA gene in duplicate. [44] Any sample with at least one positive pan *-Plasmodium* replicate was subjected to a series of four 18S rRNA real-time PCR assays specific to P. *falciparum, Plasmodium malariae, Plasmodium ovale*, and P. *vivax* in duplicate.[45–47] Species calls were only made if at least two total replicates were positive. Samples with only a single positive pan*-Plasmodium* replicate but negative species-specific assays were called negative. Samples in which both pan*-Plasmodium* replicates were positive but species-specific assays negative were subjected to a PCR assay specific to the *Plasmodium knowlesi* Pkr140 gene. [48] PCR primers and reaction conditions are described in the **Supplementary File**.

### Data analysis

We made comparisons using the Kruskal-Wallis Rank Sum or Fisher’s exact test for categorical variables and the t-test for normally distributed continuous variables. Statistical analyses were performed using R software.

## FOOTNOTES

## Data Availability

Data not contained in the manuscript or supplements is available upon reasonable request.

## Acknowledgements

The authors thank Steven Meshnick for his key role in conceptualizing this study and interpretation of results, and Nicholas Brazeau for assistance with bioinformatic pipelines and whole-genome sequencing summary statistic visualization. They also thank the study supervisors, staff, and participants. The following reagents were obtained through BEI Resources, NIAID, NIH: Genomic DNA from *P. falciparum* strain 3D7, MRA-102G, contributed by Daniel J. Carucci; *P. falciparum* strain HB3, MRA-155G, contributed by Thomas E. Wellems; *P. falciparum* strain Dd2, MRA-150G, contributed by David Walliker; and P. *knowlesi* strain H, MRA-456G, contributed by Alan W. Thomas; in addition to diagnostic plasmid containing the small subunit ribosomal RNA gene (18S) from *Plasmodium vivax*, MRA-178, *Plasmodium ovale*, MRA-180, and *Plasmodium malariae*, MRA-179, contributed by Peter A. Zimmerman.

## Presentations

Preliminary findings were presented at the American Society for Tropical Medicine and Hygiene in New Orleans, Louisiana, USA, on October 30, 2018, and at the Journées Scientifiques de Lutte contre le Paludisme 2019 conference in Kinshasa, Democratic Republic of Congo on April 30, 2019.

## Corresponding author

Jonathan B. Parr, MD, MPH, Division of Infectious Diseases, University of North Carolina, 130 Mason Farm Rd., Chapel Hill, NC 27599; phone 1-919-445-1132.

## Changes in affiliation

N/A

## SUPPLEMENTARY MATERIAL

**Supplementary Table 1.** Characteristics of study subjects by health zone.

| Province | Overall | Bas-Uele |  | Kinshasa |  | Sud-Kivu |  |
| --- | --- | --- | --- | --- | --- | --- | --- |
| Health zone |  | Buta | Ganga | Limete | Nsele | Idjwi | Kadutu |
| n | 3627 | 598 | 605 | 629 | 619 | 587 | 589 |
| Age, mean years (SD) | 20.7 (18.5) | 18.4 (17.5) | 20.6 (18.2) | 20.8 (18.8) | 20.7 (18.6) | 21.2 (18.5) | 22.9 (19.2) |
| Age strata, n (%) |  |  |  |  |  |  |  |
| <5 years | 1025 (28.7) | 177 (30.7) | 193 (32.1) | 188 (29.9) | 147 (23.7) | 179 (30.5) | 141 (25.2) |
| 5-14 years | 579 (16.2) | 107 (18.5) | 63 (10.5) | 115 (18.3) | 156 (25.2) | 51 (8.7) | 87 (15.6) |
| 15-24 years | 639 (17.9) | 116 (20.1) | 116 (19.3) | 95 (15.1) | 86 (13.9) | 133 (22.7) | 93 (16.6) |
| 25-34 years | 513 (14.4) | 79 (13.7) | 91 (15.1) | 64 (10.2) | 80 (12.9) | 107 (18.2) | 92 (16.5) |
| 35-44 years | 344 (9.6) | 34 (5.9) | 67 (11.1) | 64 (10.2) | 67 (10.8) | 50 (8.5) | 62 (11.1) |
| 45-54 years | 249 (7.0) | 37 (6.4) | 36 (6.0) | 66 (10.5) | 42 (6.8) | 31 (5.3) | 37 (6.6) |
| 55 years and older | 223 (6.2) | 27 (4.7) | 35 (5.8) | 37 (5.9) | 41 (6.6) | 36 (6.1) | 47 (8.4) |
| Female gender, n (%) | 2130 (58.7) | 403 (67.4) | 377 (62.3) | 302 (48.0) | 344 (55.6) | 342 (58.3) | 362 (61.5) |
| Pregnant, n (%) | 350 (16.4) | 81 (20.1) | 24 (6.4) | 4 (1.3) | 22 (6.4) | 119 (34.8) | 100 (27.6) |
| Slept under bednet the night before, n (%) | 2238 (79.5) | 396 (93.8) | 190 (54.3) | 480 (85.6) | 422 (76.2) | 355 (85.7) | 395 (77.0) |
| Diagnosed with malaria in the last six months, n (%) | 1556 (43.1) | 390 (65.2) | 368 (60.8) | 173 (28.3) | 289 (46.8) | 196 (33.4) | 140 (23.8) |
| Microscopy-positive, n (%) | 1397 (38.7) | 189 (31.7) | 311 (51.6) | 246 (39.5) | 287 (46.6) | 298 (50.8) | 66 (11.2) |
| RDT-positive, n (%) | 1545 (42.6) | 334 (55.9) | 424 (70.1) | 99 (15.7) | 281 (45.4) | 342 (58.3) | 65 (11.0) |
| RDT-negative, microscopy-positive, n (%) | 426 (11.8) | 14 (2.3) | 37 (6.1) | 178 (28.5) | 89 (14.4) | 69 (11.8) | 39 (6.6) |

**Supplementary Table 2.** Comparison of malaria diagnostic test results. HRP2-based RDT, *P. falciparum* 18S rRNA real-time PCR, and microscopy (micro) comparisons, counts (percent). A) RDT versus PCR. B) RDT versus microscopy. C) Microscopy versus PCR. D) RDT and PCR profiles by province.

|  |  |  |  |  |  |
| --- | --- | --- | --- | --- | --- |
| A. |  | PCR+ | PCR- |  |  |
|  | RDT+ | 404 (40) | 36 (4) |  |  |
|  | RDT- | 134 (13) | 426 (43) |  |  |
| B. |  | Micro+ | Micro- |  |  |
|  | RDT+ | 267 (27) | 173 (17) |  |  |
|  | RDT- | 108 (11) | 452 (45) |  |  |
| C. |  | PCR+ | PCR- |  |  |
|  | Micro+ | 287 (29) | 88 (9) |  |  |
|  | Micro- | 251 (25) | 374 (37) |  |  |
| D. |  | RDT-/PCR- | RDT-/PCR+ | RDT+/PCR+ | RDT+/PCR- |
| Province | Kinshasa | 234 (66) | 17 (5) | 87 (25) | 15 (4) |
|  | Sud-Kivu | 142 (45) | 54 (17) | 112 (35) | 11 (3) |
|  | Bas-Uele | 50 (15) | 63 (19) | 205 (62) | 10 (3) |

**Supplementary Figure 1.**
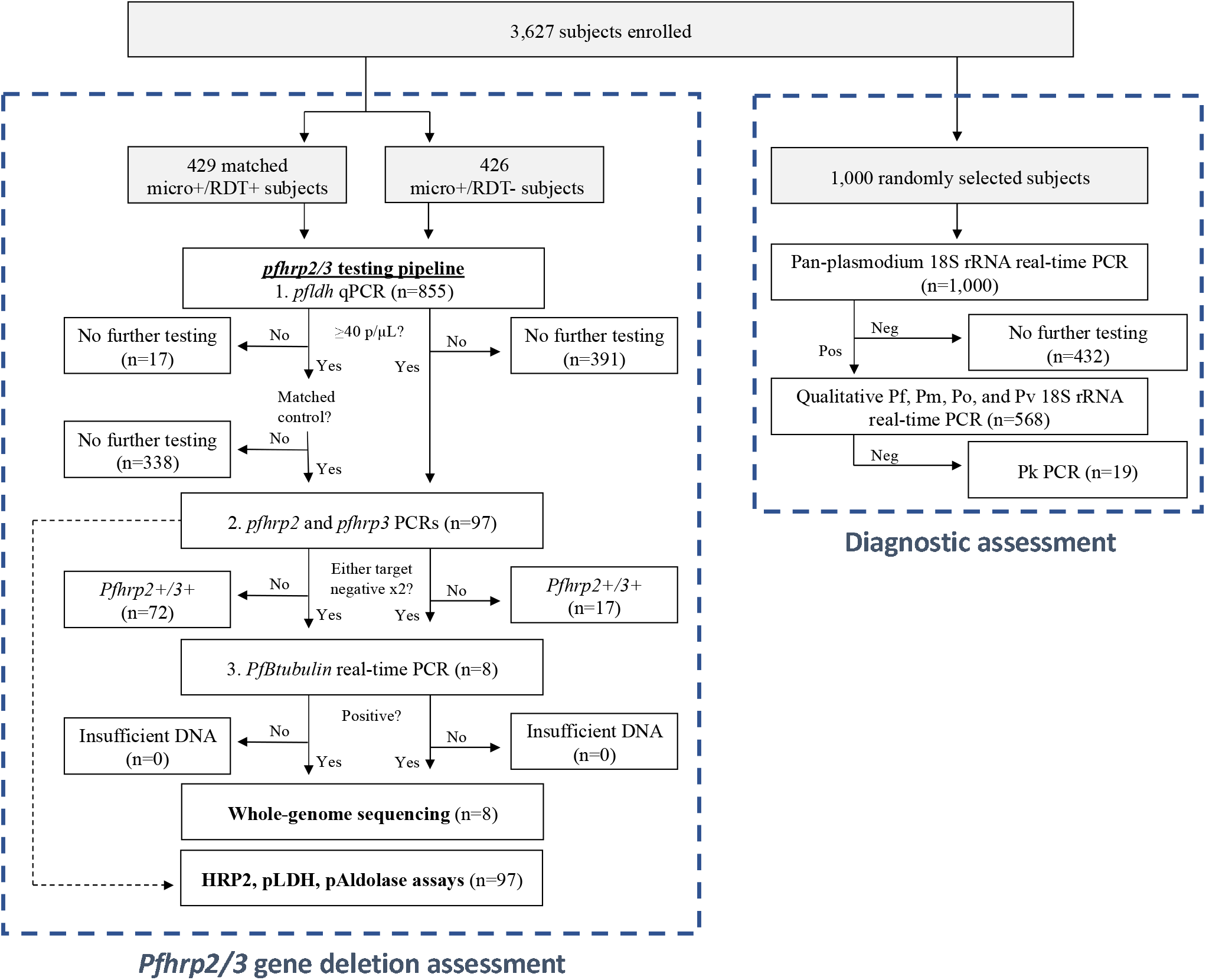
Sample selection for assessment of *pfhrp2/3* deletions and diagnostic performance. Abbreviations: Pf, *P. falciparum;* Pm, *P. malariae*; Po, *P. ovale;* Pv, *P. vivax;* Pk, *P. knowlesi*.

**Supplementary Figure 2.**
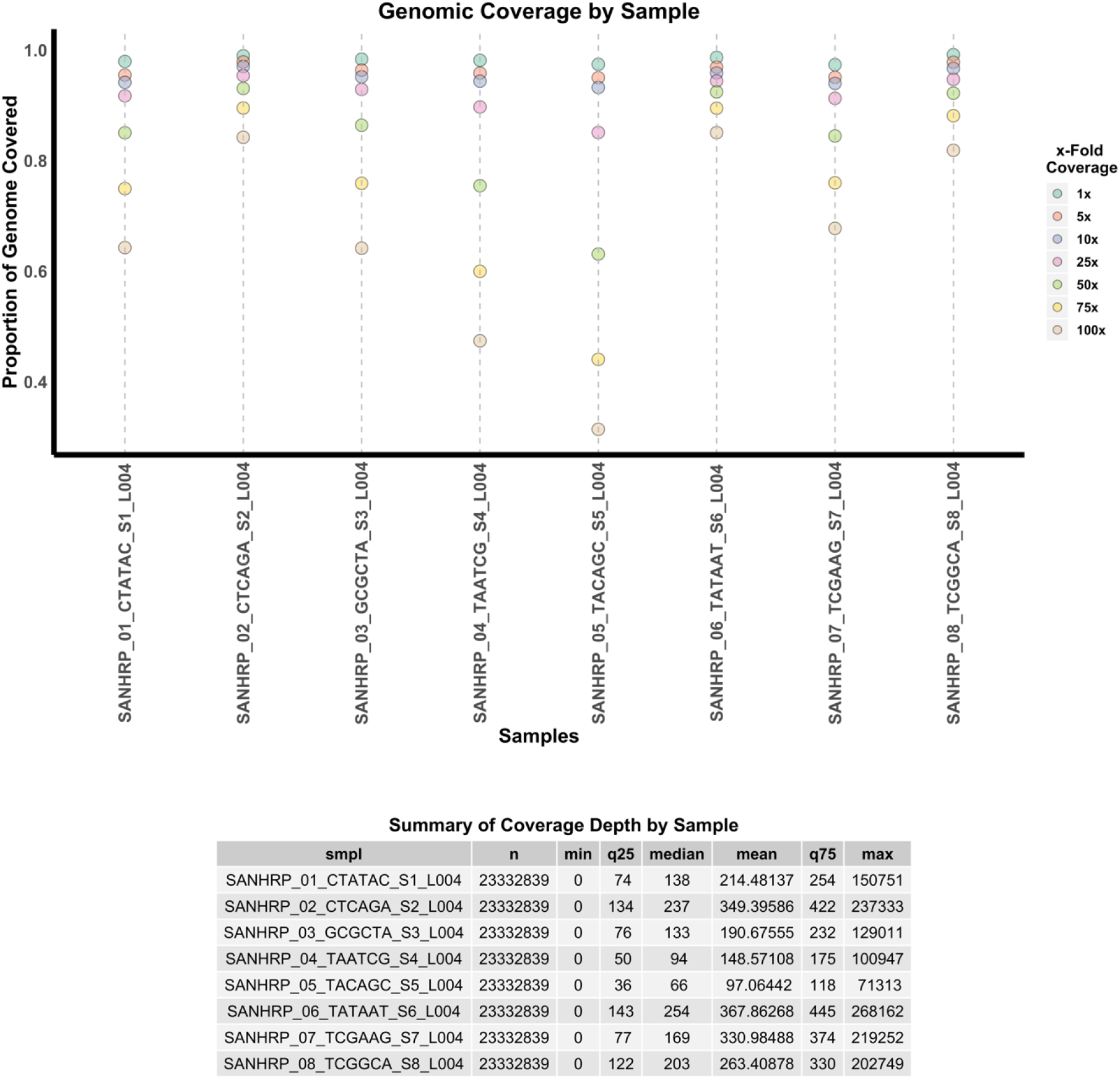
Genomic coverage statistics by sample.

**Supplementary Figure 3.**
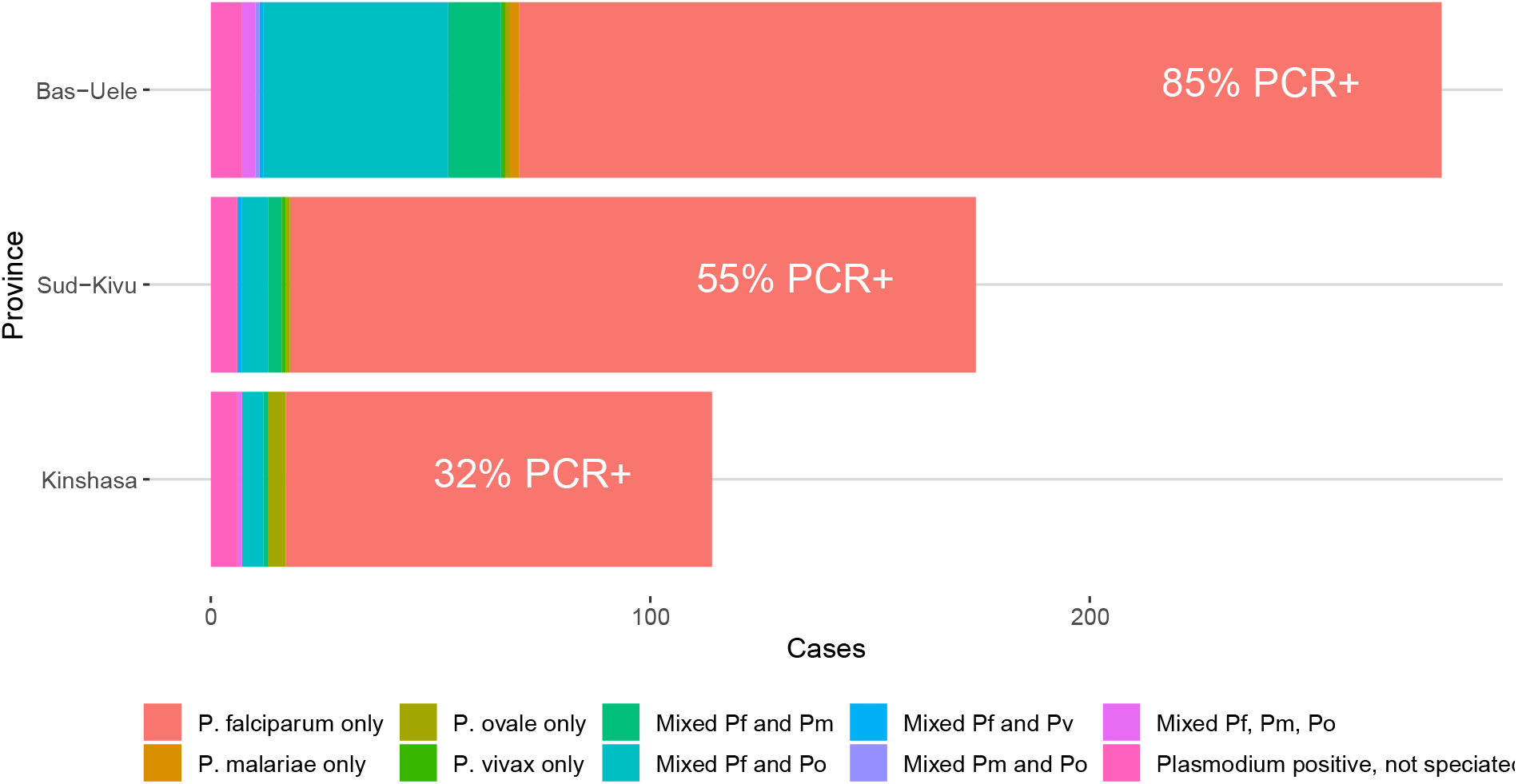
Malaria prevalence was high (57% overall) and non-falciparum co-infection common (15% overall) among symptomatic subjects. *Plasmodium* PCR prevalence and species-specific real-time PCR results for *P. falciparum, P. ovale, P. malariae*, and *P. vivax* among 1,000 samples tested, with counts reported by province. Province-level prevalence of *pan-Plasmodium* PCR-positivity is displayed in white font.

**Supplementary Figure 4.**
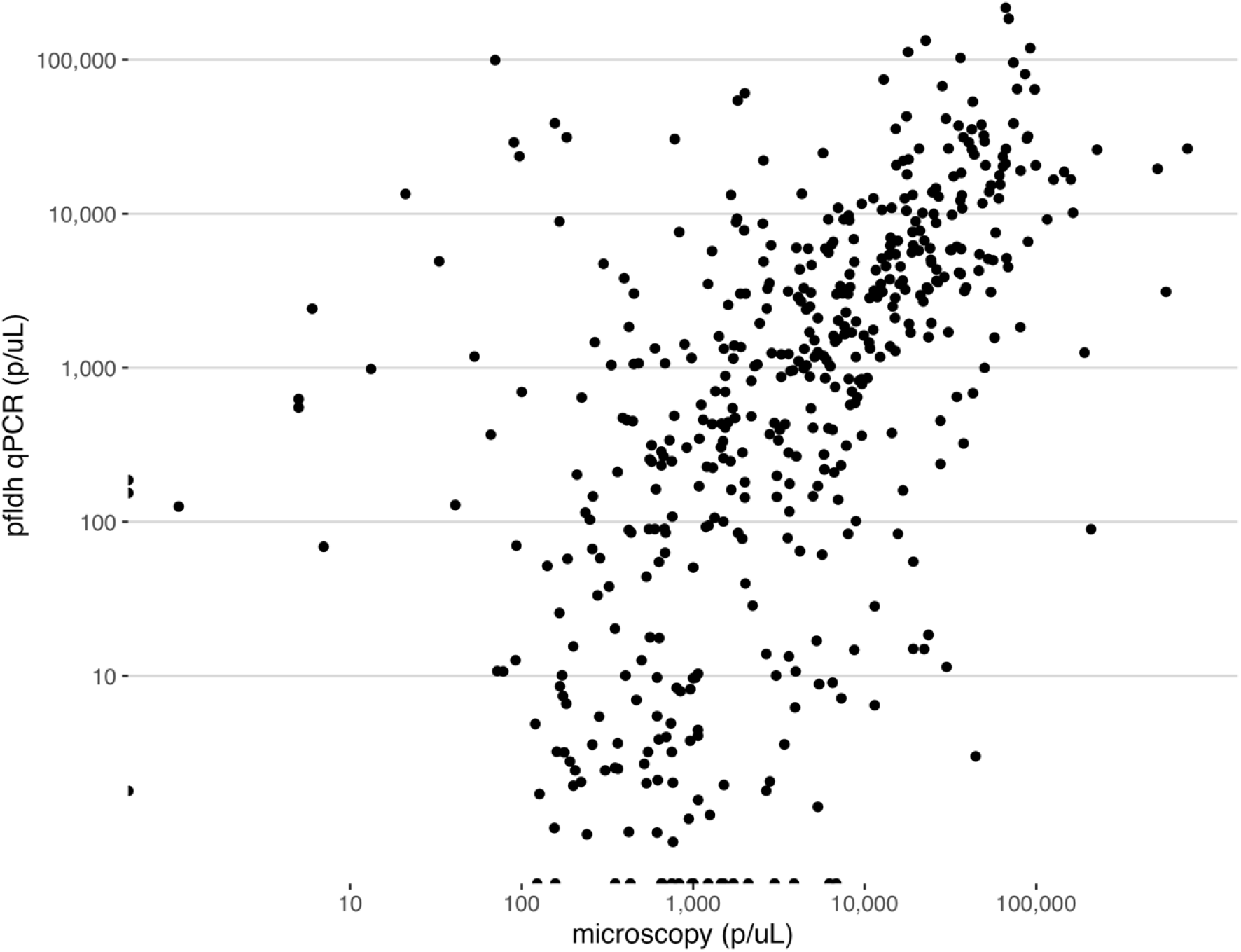
Parasite densities determined by *pfldh* qPCR and microscopy were similar, with moderate correlation (Spearman correlation coefficient: 0.63, p <0.001).

Supplementary File. PCR primers, probes, and reaction conditions.

